# Recent Medical Literature on Post-Pandemic Mental Health: An Integrative Analysis of Scientific Publications

**DOI:** 10.1101/2025.09.26.25336771

**Authors:** Bárbara Aline Ferreira Assunção

## Abstract

**Objective:** To analyze the impacts of the COVID-19 pandemic on post-pandemic mental health, identifying the most prevalent symptoms, risk factors, and coping strategies.

**Methods:** Systematic literature review conducted according to PRISMA guidelines, using open-access databases (Google Scholar and PubMed) with articles published between 2021 and 2025, in English or Portuguese, full-text available, and focused on human studies. Inclusion and exclusion criteria were applied rigorously, and data were extracted in a standardized manner for qualitative analysis of neuropsychiatric effects, psychosocial factors, and mental health coping strategies.

**Results:** A total of 26 studies were included. Depression and anxiety were the most prevalent symptoms (20–45%), while brief psychosis, acute mania, and post-COVID-19 encephalitis were less frequently reported. Risk factors included social isolation, unemployment, and comorbidities, with significant impact on children, adolescents, older adults, and healthcare professionals. Effective interventions included physical activity, psychological resilience programs, social support, and continuous clinical monitoring.

**Conclusion:** The COVID-19 pandemic had significant physical, mental, and social health impacts, particularly among vulnerable groups. Integrated strategies promoting well-being, psychological support, and clinical follow-up are essential to mitigate adverse effects. Public and institutional policies should prioritize actions targeting at-risk populations, and future research should include longitudinal studies and quantitative meta-analysis to strengthen existing evidence.

## 1. Introduction

The COVID-19 pandemic marked an unprecedented event in contemporary history, with impacts extending beyond the biological dimension of SARS-CoV-2 infection to social, economic, and psychological domains. Beginning in 2020, social isolation, human losses, and uncertainty about the future directly affected mental health on a global scale. This scenario led to a marked increase in anxiety, depression, post-traumatic stress, and other neuropsychiatric manifestations, demanding special attention from both researchers and healthcare professionals.

The primary objective of this systematic review is to analyze the impacts of the COVID-19 pandemic on post-pandemic mental health, identifying the most prevalent symptoms, risk factors, and coping strategies. The specific objectives include mapping the main psychological disorders and symptoms associated with the pandemic; examining social, clinical, and environmental factors related to symptom exacerbation; and highlighting interventions, resilience strategies, and mitigation practices reported in the literature.

The rationale for this research lies in the need to understand the enduring effects of the pandemic on mental health. Although this topic has received increasing scientific attention, the methodological diversity of studies and the rapid production of knowledge during the crisis necessitate systematic analyses that organize evidence and guide clinical practice and public policy.

The relevance of this systematic review is also supported by its role in the academic and scientific context. Unlike individual studies, systematic reviews provide a comprehensive overview of existing literature, contributing to knowledge consolidation and the identification of new research directions. Furthermore, their conclusions offer practical guidance for mental health promotion and care in the post-pandemic context, when psychological effects are likely to become more pronounced.

## 2. Theoretical Framework

### 2.1 Clinical and Physical Impacts of COVID-19

The declaration of the COVID-19 pandemic by the World Health Organization (WHO) in early 2020 marked the onset of a global crisis that extended beyond the health sphere, affecting social, economic, and political dimensions (Rodrigues, 2025). Among the measures adopted, social isolation represented an unprecedented effort by individuals, abruptly altering daily routines and lifestyles worldwide. By 2021, the virus had infected millions, leaving lasting impacts on both physical health and daily life, with repercussions that were physical and psychological in nature (Silva et al., 2025). Organs most affected included the heart, lungs, and muscles, as well as neurological and psychological systems, resulting in pain, fatigue, and significant functional limitations. Respiratory diseases were the most common manifestations, given that the lungs are the primary target of the virus, potentially leading to respiratory insufficiency, fatigue, and, in severe cases, pulmonary fibrosis.

The effects of the pandemic were also significant among vulnerable populations. Children, for example, experienced disruptions in routines that negatively affected their physical and emotional health. Studies indicate that increased sedentary behavior, prolonged screen time, and changes in dietary habits, combined with social isolation, contributed to worsening childhood obesity in vulnerable groups (Borges et al., 2025). Similarly, older adults, the population most susceptible to COVID-19, experienced a decline in quality of life due to isolation, which heightened nutritional risks and increased the need for continuous supervision to prevent malnutrition (Oliveira et al., 2025).

Clinically, SARS-CoV-2 infection presented a wide range of physical and neurological manifestations that persisted beyond the acute phase of the disease. Symptoms such as headache—often associated with pre-existing migraine episode muscle pain, and fatigue were reported in several studies (Martins et al., 2025). Headaches were particularly prevalent, especially among women and individuals with a history of migraines and could persist after acute infection. Furthermore, patients with pre-existing neurological or cognitive conditions faced additional challenges in diagnosis and treatment, as exemplified in cases of progressive dementia, where overlapping COVID-19-related symptoms complicated clinical evaluation (Tayyebi et al., 2022).

Beyond physical impacts, the COVID-19 pandemic revealed significant neuropsychiatric consequences. Sprenger et al. (2022) reported that SARS-CoV-2 infection may be associated with psychosis, neurocognitive disorders, and mood disturbances, even in patients without prior psychiatric history. A clinical case illustrated the manifestation of acute mania following COVID-19 infection, highlighting that factors such as stress, grief, and anxiety related to social isolation can exacerbate neuropsychiatric symptoms. This case reinforces the need for post-infection psychiatric evaluation, considering the virus’s neurotropic properties and associated systemic inflammation.

### 2.2 Impacts on Mental Health and Psychological Well-Being

Additional studies highlight the complexity of neuropsychiatric diagnosis during the pandemic. Faisal et al. (2021) demonstrated that psychotic symptoms may emerge within days of infection, associated with laboratory alterations such as elevated D-dimer and fibrinogen levels, suggesting microlesions in the central nervous system. Kozato, Mishra, and Firdosi (2021) reported a case of severe anxiety and delusions in a patient without prior psychiatric history, emphasizing the importance of early identification and appropriate management of neuropsychiatric complications in acute hospital settings.

Furthermore, Poluga et al. (2025) documented cases of encephalitis in COVID-19 patients, even when initial radiological and CT examinations showed no significant alterations. Such manifestations, diagnosed through exclusion of other etiological agents, illustrate the diagnostic challenge and the need for comprehensive therapeutic approaches, including anti-inflammatory agents, anticoagulants, and symptomatic support. Ziemele et al. (2021) described a fatal case of acute necrotizing encephalopathy (ANE) associated with COVID-19, suggesting immune-mediated mechanisms in its pathogenesis and highlighting the importance of early diagnosis and intervention with immunomodulators.

Even patients with mild or asymptomatic infection may experience persistent neuropsychiatric complications. Jozuka et al. (2022) reported a 55-year-old woman who developed delirium and post-COVID-19 encephalopathy during home quarantine, demonstrating that mild respiratory symptoms do not preclude neurological involvement. Proper clinical monitoring and therapeutic management are essential to reduce sequelae and improve patients’ quality of life.

The literature also emphasizes psychological and emotional impacts on non-infected populations. Melo Junior et al. (2024) highlights psychological resilience as an adaptive factor, influenced by social support, coping strategies, and psychological interventions, mitigating anxiety, depression, and stress. Among healthcare professionals, Castro and Wolff Filho (2024) identified a high prevalence of burnout, anxiety, and stress-related disorders, exacerbated by workload, exposure to suffering, and stigmatization of emotional care. Psychological support strategies, resilience training, and practices such as mindfulness were found effective, although institutional barriers and stigma continue to hinder systematic implementation.

Borges (2024) notes that the two most prevalent mental health disorders in Brazil are anxiety and depression, positioning the Brazilian population among the most anxious in the world and sixth in depression prevalence. This situation is particularly concerning among young adults and health students, where high academic stress contributes to depressive symptoms, even in individuals who previously had lower baseline levels prior to entering higher education.

### 2.3 Protective Factors, Adaptation, and Mitigation Strategies

Barriers to public health and prevention were also reflected in the acceptance of preventive measures, such as vaccination. Menezes Junior et al. (2024) highlight that factors such as adverse effects, doubts about efficacy, and concerns about multiple doses negatively impacted vaccination campaigns, exacerbating social vulnerabilities during the pandemic.

In a global context, Chen et al. (2021) observed a high prevalence of mental health symptoms among healthcare workers and the general population in Africa, attributed to fear, uncertainty, and pandemic containment strategies. Pre-existing conditions, including oxygen shortages, multimorbidity, low income, and inadequate healthcare infrastructure, amplified the impact on mental health, revealing regional particularities in coping with COVID-19. França and Costa (2024) also identified challenges faced by healthcare workers in vulnerable areas, such as Endemic Agents in rural Alagoas, whose routines and habits required adaptation to preventive measures and to safeguarding both personal and public health.

Within nursing, Souza (2024) emphasizes that mental health involves emotional balance and the ability to cope with daily challenges. Nurses faced continuous exposure to stress and suffering, intensified by technology use, competitiveness, and ambitious targets, highlighting the need for policies and strategies focused on the psychological well-being of these professionals.

Virus detection and monitoring also influenced mental health and risk perception. Velozo and Peixoto (2021) underscore the importance of accurate COVID-19 diagnosis, highlighting Real-Time RT-PCR as an effective technique for identifying viral genetic material, ensuring precision amid uncertainty and fear. Ahmed (2023) reinforces that, although mental health problems increased during the pandemic, the utilization of mental health services initially declined, with gradual recovery throughout 2020 and 2021, yet in some services, levels did not return to pre-pandemic baselines.

Munindradasa et al. (2021) describe the pandemic as a “perfect storm” for mental health deterioration, capable of exacerbating pre-existing conditions or triggering new psychological problems. In extreme cases, Nardi et al. (2022) documented suicidal tendencies, including suicide pacts, exacerbated by economic difficulties and fear of infection, highlighting individual vulnerability factors, such as traits on the autism spectrum, which increase the risk of severe psychopathological consequences following traumatic events.

The pandemic’s impact on lifestyle habits is also closely related to mental health. Li, Wang, and Shen (2022) demonstrated that regular high-intensity physical activity lasting 30 to 60 minutes daily was associated with reduced anxiety, depression, and negative emotions, whereas physical inactivity increased the risk of psychological disorders and lower subjective well-being. Hammon, Pinheiro, and Patrício (2024) observed that strategies implemented by professionals from the Expanded Family Health and Primary Care Team (NASF-AB) were crucial in promoting physical activity during the pandemic, reinforcing the importance of ongoing health promotion and prevention efforts, even under social restrictions.

## 3. Methods

This study is characterized as a systematic literature review, conducted in accordance with the PRISMA (Preferred Reporting Items for Systematic Reviews and Meta-Analyses) guidelines, ensuring transparency, reproducibility, and methodological rigor throughout all stages of the review. The objective was to identify and analyze recent publications on post-pandemic mental health following COVID-19, with a focus on symptoms, risk factors, social impact, and interventions.

### 3.1 Databases and Search Strategy

Open-access electronic databases, including Google Scholar and PubMed, were used. The search was conducted according to the following criteria: articles published between 2021 and 2025, in Portuguese or English, available in full text, and focused on human studies. The search strategy involved combinations of specific terms related to COVID-19 and mental health, including but not limited to: “COVID-19,” “SARS-CoV-2,” “mental health,” “psychological impact,” “depression,” and “anxiety.” Filters for language, publication year, article type, and full-text availability were applied to ensure the inclusion of studies relevant to the research topic.

### 3.2 Inclusion and Exclusion Criteria

#### Inclusion Criteria

The inclusion criteria considered studies published between 2021 and 2025, written in English or Portuguese, and available in full text under open access. Only studies conducted with human subjects and directly addressing the relationship between mental health and the COVID-19 pandemic were included.

#### Exclusion Criteria

Studies conducted with animals, texts not available in full access, publications outside the established period, and non-systematic reviews or reports not directly related to the proposed theme were excluded.

### 3.3 Selection Process and Data Extraction

The selection of articles was carried out on different stages and across multiple databases. In Google Scholar, the initial search retrieved 2,910,000 results. After applying filters related to publication period, language, and article type, 478 studies were selected. Title screening reduced this number to 67, and the final critical appraisal resulted in the inclusion of 17 articles. In PubMed, the initial search yielded 41,958 results. Following the application of temporal, language, article type, human population, and age group filters, 121 studies remained. Title screening reduced this number to 29, and subsequent duplicate removal and critical appraisal led to the final inclusion of 9 articles.

Data extraction was performed in a standardized manner, considering information on the study population, methodology, main findings, and relevant conclusions. The analysis was conducted qualitatively, allowing the integration and synthesis of evidence, with emphasis on neuropsychiatric effects, psychosocial factors, and mental health coping strategies in the post-pandemic period.

## 4. Results

### 4.1 Study Selection

The article selection process followed the PRISMA flow (Figure 1). A total of 2,910,000 records were identified in Google Scholar and 41,958 in PubMed (Table 1). After applying filters for publication year, language, article type, and full-text availability, as well as screening titles and abstracts, the following results were obtained:

**Table 1.** Study Selection.

| Database | Initial Records | After Filters | After Title Screening | After Critical Review & Duplicate Removal | Included Articles |
| --- | --- | --- | --- | --- | --- |
| Google Scholar | 2,910,000 | 478 | 67 | 17 | 17 |
| PubMed | 41,958 | 121 | 29 | 9 | 9 |
| Total |  |  |  |  | 26 |

Source: Research data, 2025

**Figure 1.**
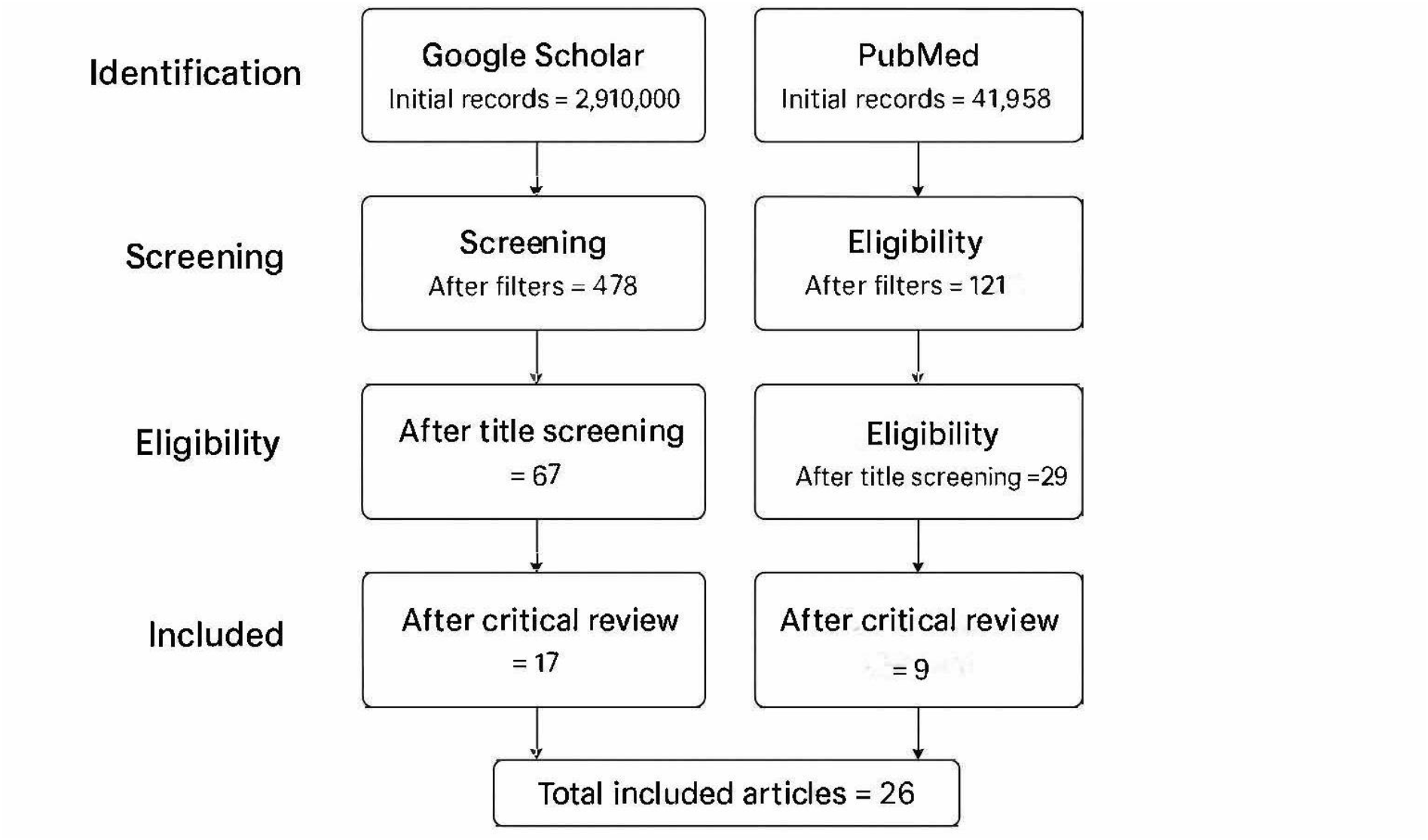
PRISMA Flow Diagram. ***Source:*** *Research data, 2025*

### 4.2 Main Findings

#### Prevalence of Mental Health Symptoms

Depression and anxiety were the most reported symptoms across different populations, ranging from 20% to 45% depending on the region and study group (Ahmed et al., 2023; Li et al., 2022). Cases of brief psychosis and acute mania post-COVID-19 were documented in patients with no prior psychiatric history (Faisal et al., 2021; Sprenger et al., 2022; Kozato et al., 2021). Encephalitis associated with SARS-CoV-2 was identified in severe case reports (Poluga et al., 2025; Tayyebi et al., 2022; Ziemele et al., 2021).

#### Risk Factors and Social Impact

Social isolation, unemployment, and comorbidities increased the risk of mental health symptoms (Rodrigues, 2025; Silva et al., 2025). Children and adolescents showed increased obesity and emotional problems, associated with confinement and school interruption (Borges et al., 2025). Healthcare professionals reported high stress, burnout, and emotional challenges during the pandemic (Souza, 2024; Castro & Wolff Filho, 2024; Menezes Junior et al., 2024).

#### Interventions and Coping Strategies

Physical activity showed a positive effect in reducing anxiety and depression (Li et al., 2022; Hammon et al., 2024). Psychological resilience programs and social support were associated with better emotional outcomes (Melo Júnior et al., 2024; Borges, 2024). Monitoring and management by primary healthcare professionals were essential to identify and refer severe cases (Munindradasa et al., 2021).

Below, Table 2 presents the characteristics of the included studies, and Table 3 details the types of post-COVID-19 neuropsychiatric symptoms identified in the selected research.

**Table 2.** Characteristics of the included studies.

| Author(s) | Year | Country/Region | Study type | Population | Main findings |
| --- | --- | --- | --- | --- | --- |
| Ahmed et al. | 2023 | Europe | Systematic review | General | High prevalence of anxiety and depression; significant social impact |
| Li et al. | 2022 | China | Systematic review | General population | Physical activity reduces depression and anxiety symptoms |
| Faisal et al. | 2021 | Indonesia | Case report | Patient with COVID-19 | Brief psychosis in a patient with no prior history |
| Poluga et al. | 2025 | Serbia | Case report | Patients with COVID-19 | Encephalitis associated with SARS-CoV-2 |
| Rodrigues | 2025 | Brazil | Article | General population | Social and psychological impact; critical social isolation |
| Souza | 2024 | Brazil | Integrative review | Nursing professionals | High levels of burnout; need for institutional support |
| Borges et al. | 2025 | Brazil | Article | Children/adolescents | Childhood obesity and emotional impact in the post-pandemic period |
| Hammon et al. | 2024 | Brazil | Integrative review | General population | Physical activity interventions as coping strategies |

*Source: research data, 2025.*

**Table 3.**
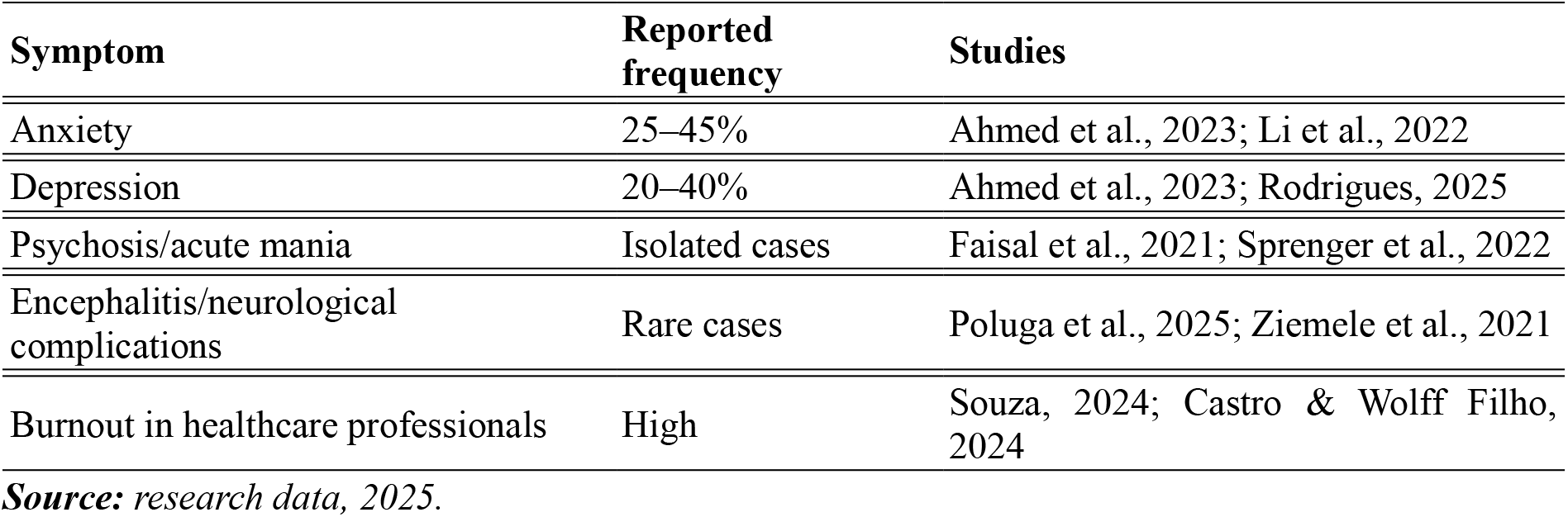
Types of post-COVID-19 neuropsychiatric symptoms.

## 5. Discussion

### 5.1 Interpretation of Results

The findings of this systematic review demonstrate that the COVID-19 pandemic had significant impacts on the physical and mental health of the populations studied. Symptoms of anxiety and depression were the most prevalent, affecting between 20% and 45% of individuals (Ahmed et al., 2023; Li et al., 2022), supporting the notion that social isolation and fear of infection generated widespread psychological distress. Brief psychosis, acute mania, and encephalitis associated with infection, although rare, highlight the neurotropism of the virus and the effects of systemic inflammation on the central nervous system (Faisal et al., 2021; Poluga et al., 2025; Sprenger et al., 2022; Tayyebi et al., 2022).

Physical impacts were also relevant, with fatigue, headache, and respiratory limitations being particularly noteworthy. Children and adolescents experienced increased obesity, sedentary behavior, and emotional changes, while older adults showed heightened vulnerability due to malnutrition and social isolation (Borges et al., 2025; Oliveira et al., 2025). These data suggest that the effects of the pandemic went beyond the acute disease, affecting both infected individuals and uninfected populations in socially vulnerable contexts.

### 5.2 Comparison with Previous Literature

Our results are consistent with international studies reporting a high prevalence of anxiety and depression in the post-pandemic period (Ahmed et al., 2023; Chen et al., 2021). The occurrence of neurological complications, such as encephalitis and post-COVID-19 delirium, has also been documented in recent reviews (Ziemele et al., 2021; Jozuka et al., 2022), reinforcing that even mild infections can lead to long-lasting neuropsychiatric symptoms.

In the Brazilian context, local literature corroborates the psychological impact on healthcare professionals and students, highlighting elevated stress, burnout, and increased depression and anxiety (Castro & Wolff Filho, 2024; Borges, 2024). Moreover, interventions such as physical activity, resilience strategies, and social support were effective in mitigating adverse effects, confirming findings reported by Li et al. (2022) and Hammon et al. (2024).

### 5.3 Study Limitations

Despite methodological rigor, some limitations should be considered. The review included only articles in Portuguese and English, which may have excluded relevant studies published in other languages. Additionally, the heterogeneity of methods and study populations makes direct comparison of findings difficult. Case reports and case series constitute part of the analyzed material, limiting the generalizability of the results. Finally, the review did not perform a quantitative meta-analysis, being restricted to a qualitative synthesis.

### 5.4 Practical Implications

The results of this review have important clinical and social implications. Healthcare professionals should consider psychiatric and neurological follow-up for post-COVID-19 patients, even for those who experienced mild infection. Health promotion strategies, including encouragement of physical activity, psychological support, and resilience programs, are essential to mitigate psychological impacts and improve quality of life.

Public policies should prioritize interventions targeted at vulnerable groups, such as children, adolescents, and older adults, ensuring access to nutritional care, emotional support, and health education. Furthermore, institutional programs aimed at healthcare professionals can help reduce burnout and enhance emotional well-being during health crises.

## Final Considerations

This systematic review demonstrated that the COVID-19 pandemic significantly impacted the physical, mental, and social health of the populations studied. A high prevalence of anxiety and depression was observed, as well as rare neuropsychiatric manifestations such as psychosis, acute mania, and encephalitis, in addition to persistent physical repercussions including fatigue, headache, and respiratory limitations. Children, adolescents, and older adults were shown to be particularly vulnerable, presenting routine disruptions, increased sedentary behavior, obesity, and nutritional impairment. Healthcare professionals and students in demanding academic programs reported high stress levels, burnout, and emotional disorders, highlighting the wide-ranging effects of the pandemic across different population groups.

The findings of this review also emphasize the importance of mitigation strategies, such as regular physical activity, psychological resilience programs, social support, and continuous clinical monitoring, which proved effective in reducing symptoms and improving well-being. These results reinforce the need for integrated interventions that address both the physical and psychological dimensions of post-pandemic health. Moreover, institutional and public policies should prioritize the protection of vulnerable groups by ensuring access to healthcare, emotional support, and health education.

For clinical practice, neurological and psychiatric follow-up of post-COVID-19 patients is essential, even for those with mild or asymptomatic infections, along with the promotion of healthy lifestyle habits that support physical and mental well-being. The creation of institutional programs providing emotional support to healthcare professionals is also crucial to prevent burnout and promote emotional balance in the face of health crises. In terms of future research, longitudinal studies are recommended to investigate the persistence of physical and neuropsychiatric symptoms across age groups, as well as the evaluation of specific interventions such as physical exercise, psychological support, and health education. Expanding analyses to diverse populations, including studies in other languages, and conducting quantitative meta-analyses are essential to strengthen the available evidence.

In summary, the COVID-19 pandemic underscored the need for integrated approaches to promote physical, mental, and social health. The combination of clinical follow-up, appropriate public policies, and strategies to foster well-being proves vital in reducing adverse impacts, supporting population adaptation, and strengthening resilience in the face of global health crises.

## Data Availability

All data produced in the present work are contained in the manuscript

